# Quantitative Evaluation of apparent diffusion coefficient in a large multi-unit institution using the QIBA diffusion phantom

**DOI:** 10.1101/2020.09.09.20191403

**Authors:** Joshua P Yung, Yao Ding, Ken-Pin Hwang, Carlos E. Cardenas, Hua Ai, Clifton David Fuller, R. Jason Stafford

## Abstract

**Purpose:** The purpose of this study was to determine the quantitative variability of diffusion weighted imaging and apparent diffusion coefficient values across a large fleet of MR systems. Using a NIST traceable magnetic resonance imaging diffusion phantom, imaging was reproducible and the measurements were quantitatively compared to known values.

**Methods:** A fleet of 23 clinical MRI scanners was investigated in this study. A NIST/QIBA DWI phantom was imaged with protocols provided with the phantom. The resulting images were analyzed and ADC maps were generated. User-directed region-of-interests on each of the different vials provided ADC measurements among a wide range of known ADC values.

**Results:** Three diffusion phantoms were used in this study and compared to one another. From the one-way analysis of the variance, the mean and standard deviation of the percent errors from each phantom were not significantly different from one another. The low ADC vials showed larger errors and variation and appear directly related to SNR. Across all the MR systems and data, the coefficient of variation was calculated and Bland-Altman analysis was performed. ADC measurements were similar to one another except for the vials with the lower ADC values, which had a higher coefficient of variation.

**Conclusion:** ADC values among the three phantoms showed good agreement and were not significantly different from one another. The large percent errors seen primarily at the low ADC values were shown to be a consequence of the SNR dependence and very little bias was observed between magnetic strengths and manufacturers. ADC values between diffusion phantoms were not statistically significant. Future investigations will be performed to study differences in magnetic field strength, vendor, MR system models, gradients, and bore size. More data across different MR platforms would facilitate quantitative measurements for multi-platform and multi-site imaging studies. With the increasing usage of diffusion weighted imaging in the clinic, the characterization of ADC variability for MR systems provides an improved quality control over the MR systems.

## Introduction

Diffusion weighted imaging (DWI) is increasingly being used in oncologic applications for assessment of disease response and prediction of therapeutic effects [1–9]. In head and neck cancers, in particular, there is a growing body of literature, which supports, at least in preliminary institutional datasets, the utility of DWI for prognostication of radiotherapeutic or chemoradiotherapeutic response [10–13].

However, the means for assessing the standardization of apparent diffusion coefficient (ADC) measurements across individual MRI scanners has, until recently, been an unmet need. While apparent diffusion coefficient measurements represent an actual physical parameter, the variance across inter-institutional, inter-device, field strength dependence, and acquisition dependencies have yet to be fully characterized. As DWI is more frequently employed as a quantitative imaging biomarker, harmonization across devices, manufacturers, field strengths and acquisition parameters is necessary to ensure transferability and scalability of large scale clinical trials and datasets.

A sampling of 23 clinical MRI scanners was investigated in this study. DWI acquisitions with ADC reporting are performed in all of these scanners allowing for patients to be scheduled for iterative evaluation and serial imaging across different device manufacturers, models, field strengths, and acquisition parameters. In order to assess harmonization and standardization across the fleet for developing, implementing and continuously assessing a quality control program for quantitative biomarker imaging we provide an initial characterization of the variance in our institutional DWI and ADC performance across our fleet using a newly constructed jointly developed NIST/QIBA DWI phantoms (High Precision Devices Inc., Boulder, CO, USA). We also hope to utilize this information to categorize our scanners by vendor, platform, or field strength by those which are closest in terms of ADC accuracy, so that, if necessary, patients can be selectively triaged to scanners which most closely approximate the machine of their initial imaging.

This initial benchmarking of the scanners using the QIBA protocol is the precursor to longitudinal assessment amongst scanners for quality assurance as well as later lending to the investigation of variance introduced by specific clinical protocols and advanced acquisition techniques (e.g., multi-shot, simultaneous multi-slice encoding, etc).

## Materials and Methods

In this study, three NIST/QIBA phantoms were evaluated on 23 MRI systems constituting nine platforms (Table 1). A total of four 1.5T platforms and four 3T platforms were investigated. Four platforms were Siemens (Siemens Healthcare, Erlangen, Germany) and five were GE (GE Medical Systems, Milwaukee, WI, USA). Note the 3.0T GE Signa HDxt systems had Twin Resonance Module (TRM) gradients which allowed the acquisitions to be performed in two separate performance modes.

**Table 1:**
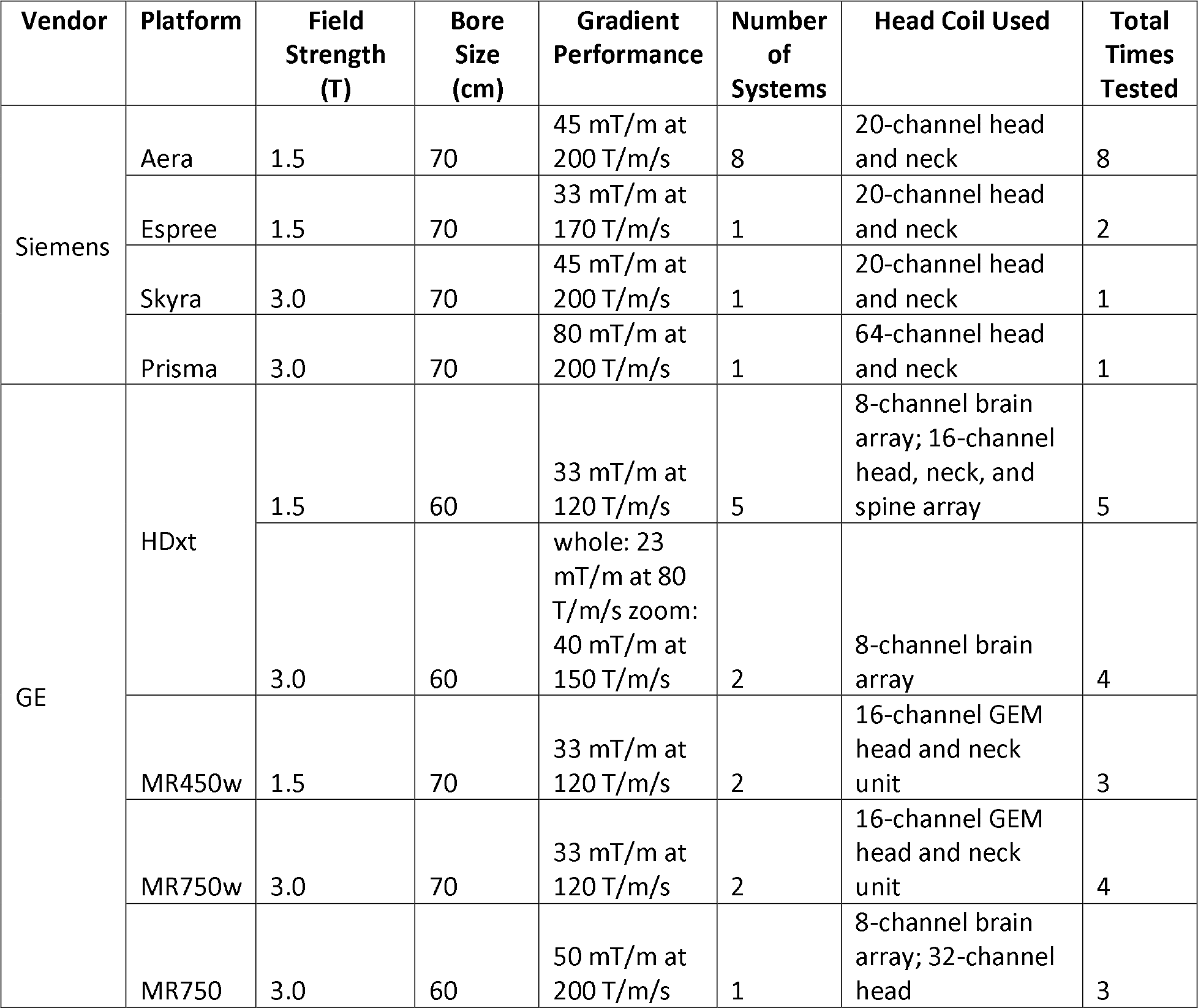
List of MRI systems.

The NIST-traceable QIBA diffusion phantom [14] is 194mm in diameter and holds 30mL vials of polymer polyvinylpyrrolidone (PVP) in aqueous solution. The vials have a range of concentrations of PVP allowing for a range of ADC values from 118 to 1091 ×10^-6^ mm^2^/s. Increasing concentrations of the polymer cause the ADC values to be reduced. The 13 vials within the phantom consist of three vials of pure water and two vials each of a PVP solution at 10, 20, 30, 40, 50% PVP by mass fraction. To eliminate the temperature sensitivity of diffusion, the phantom was filled with ice water prior to scanning to equilibrate the phantom at 0°C according to the provided phantom instructions [14]. The value of the diffusion at 0°C as a function of PVP is given in the table below (Table 2).

**Table 2:** ADC values of each vial at 0°C given by the manufacturer. 0c refers to the 0% PVP concentration vial in the center. Each PVP concentration level has an inner and outer vial represented by a ‘i’ or ‘o’ suffix in the table. For example, 0i and 0o refer to the inner and outer 0% PVP concentration vials, respectively.

| Concentration (%) | References ($\times 10^{-6} \text{ mm}^2/\text{s}$) |
| --- | --- |
| 0c | 1091.060571 |
| 0i | 1090.134463 |
| 0o | 1090.21889 |
| 10i | 824.0288105 |
| 10o | 824.1549136 |
| 20i | 596.9578014 |
| 20o | 599.359388 |
| 30i | 393.699701 |
| 30o | 396.467332 |
| 40i | 235.1549486 |
| 40o | 238.4391252 |
| 50i | 118.275699 |
| 50o | 122.9606508 |

The diffusion weighted acquisition parameters followed the recommended QIBA protocol, but had some platform dependent parameters. The phantom was imaged using the system’s head phased-array radiofrequency receive coil which ranged from 8-64 channels (see Table 1). Imaging was performed using diffusion-weighted, single-shot echo planar imaging (EPI) acquisitions (TE = 71.8-113ms; TR = 10 or 15s; Field of view = 216-220mm; Matrix = 128×128 or 192×192; Slice thickness = 4mm; Number of averages = 1) with four b-values (0, 500, 900, 2000 s/mm^2^). The NIST/QIBA phantoms were first characterized to establish phantom equivalence via analysis of their variance. Subsequently, each scanner was evaluated. Scanners with dual gradient modes (n=2) were evaluated in each mode.

The resulting images were analyzed software developed within the laboratory of Dr. Chenevert that generated ADC maps from the four b-values with a user-directed region-of-interest (ROI) placement (Figure 2). The quantitative measurements were compared to the NIST-measured ADC values provided by the phantom manufacturer. The coefficient of variation (CoV) was calculated for each PVP concentration and slice orientation: axial, coronal, and sagittal. The dependency of the ADC accuracy to the signal-to-noise ratio (SNR) of the image was also examined. Bland-Altman plots and their respective limits of agreement [15, 16] were used to investigate the ADC variability and bias between devices, manufacturer, magnetic field strength.

**Figure 1:**
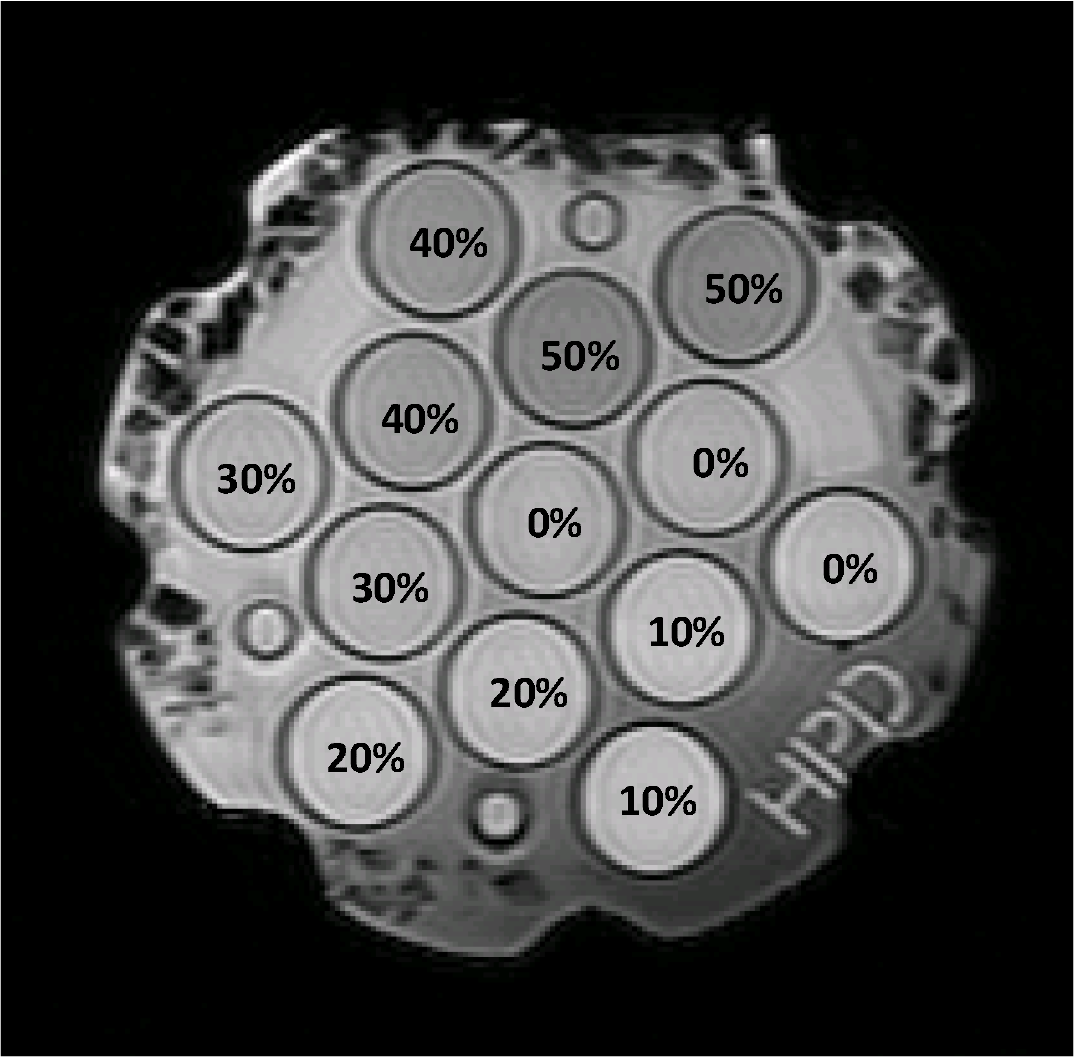
A MR image showing the 30 mL vials of aqueous solutions of PVP. Each solution has an inner and outer vial. The measured values are listed in Table 2.

**Figure 2:**
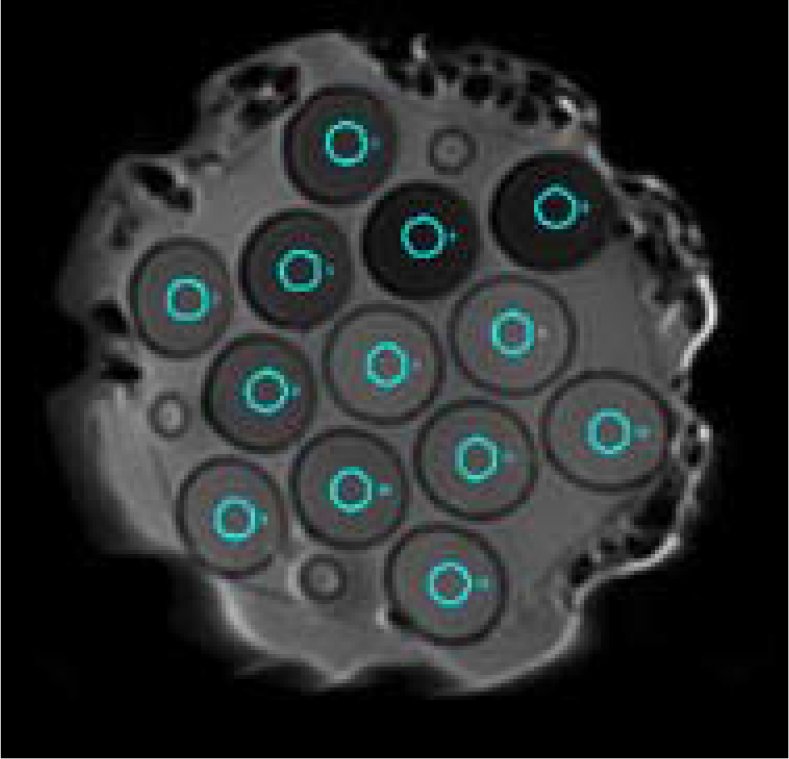
Typical example of diffusion-weighted image analysis showing the user-selected region of interests (cyan) overlaid on to each of the 13 internal vials. ROI’s were sized and placed in a manner to avoid partial volume effects with the vial edge.

## Results

In total, 23 different MR systems were investigated in this study. Including the two systems with dual gradient modes and some scanners being scanned multiple times across the three phantoms to validate the equivalence of the phantoms, a total of 31 sets of data were collected.

In Figure 3, a comparison of the three diffusion phantoms used in this study is shown. Each phantom was scanned on the same MR system and the percent error of the ADC measurement compared to the known value was calculated. The percent error was consistently less than 10% except for the 50% PVP concentration which corresponded to the lowest ADC value (118.3-123.0 ×10^-6^ mm^2^/s). Standard deviation also increased predictably as the ADC values decreased. In the one-way analysis of the variance (ANOVA) the ADC values between the diffusion phantoms were not statistically significant (10% PVP: F = 0.34, p = 0.72, 20% PVP: F = 0.88, p = 0.44, 30% PVP: F=0.01, p = 0.99, 40% PVP: F=0.31, p = 0.74, 50% PVP: F = 0.22, p=0.80) except for the water references (0% PVP: F=10.3, p>0.05), where the errors were the smallest. The large increase in error and standard deviation appear to be directly related to the SNR (Figure 4).

**Figure 3:**
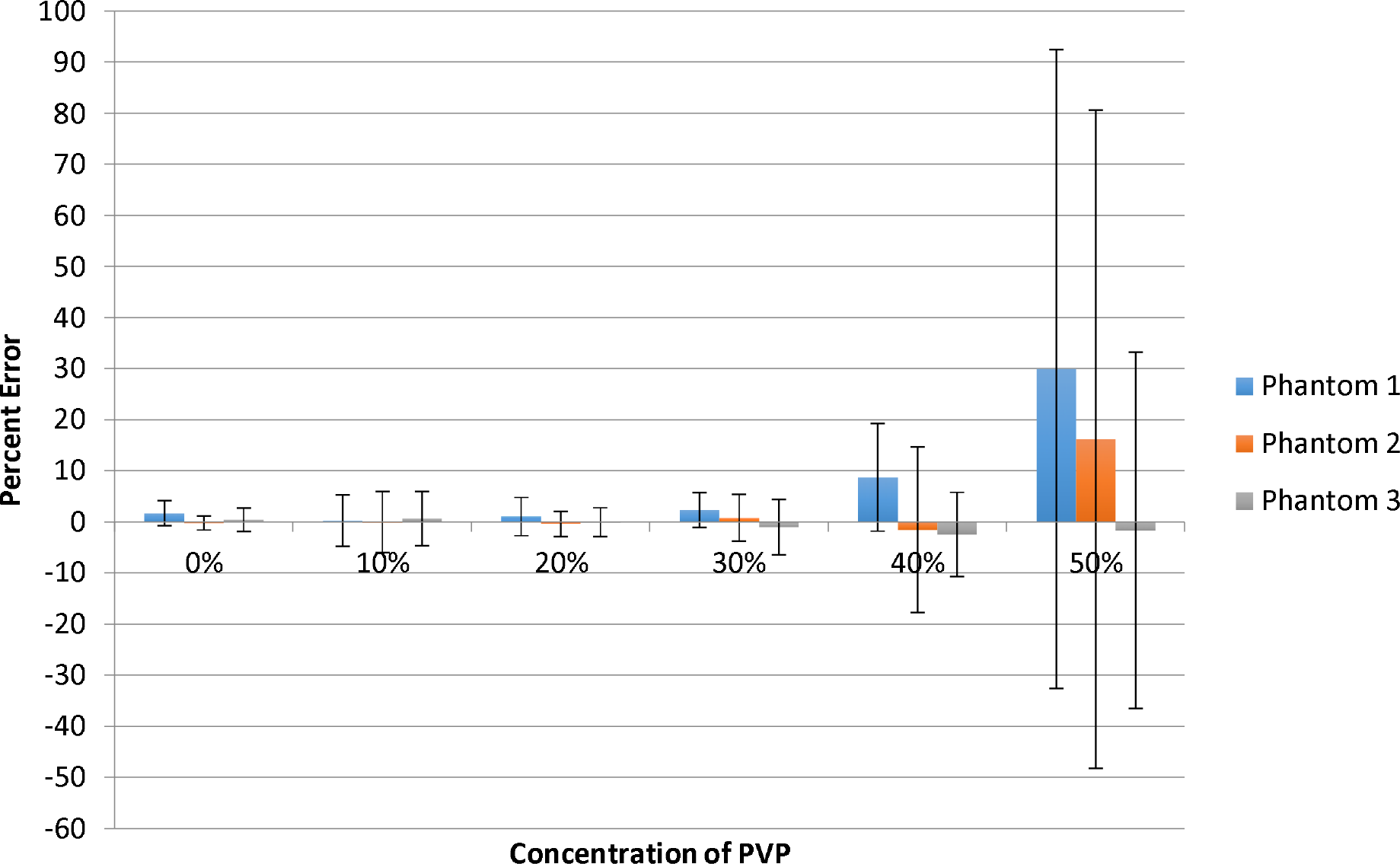
The average percent error for each concentration of PVP in one MR system that scanned all three diffusion phantoms.

**Figure 4:**
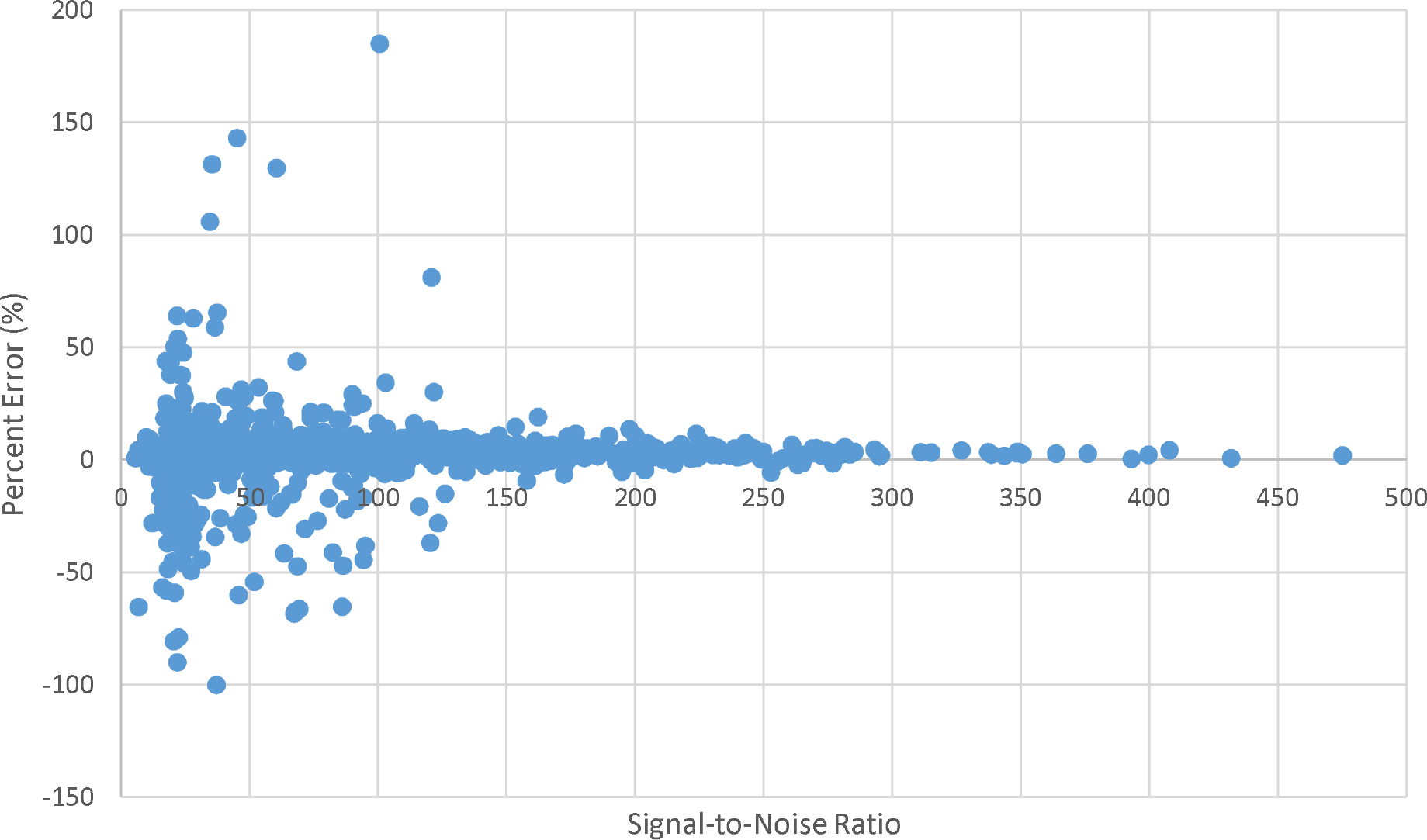
Scatter plot of the software-measured signal-to-noise ratio versus the percent error in the measurement.

The coefficient of variation (CoV) was calculated for each dataset with respect to the PVP concentration and slice orientation (Figure 5–6). Across all the MR systems and dataset, the vials with the higher PVP concentrations had the higher CoV. The sagittal slice orientation also had the highest CoV among the different orientations.

**Figure 5:**
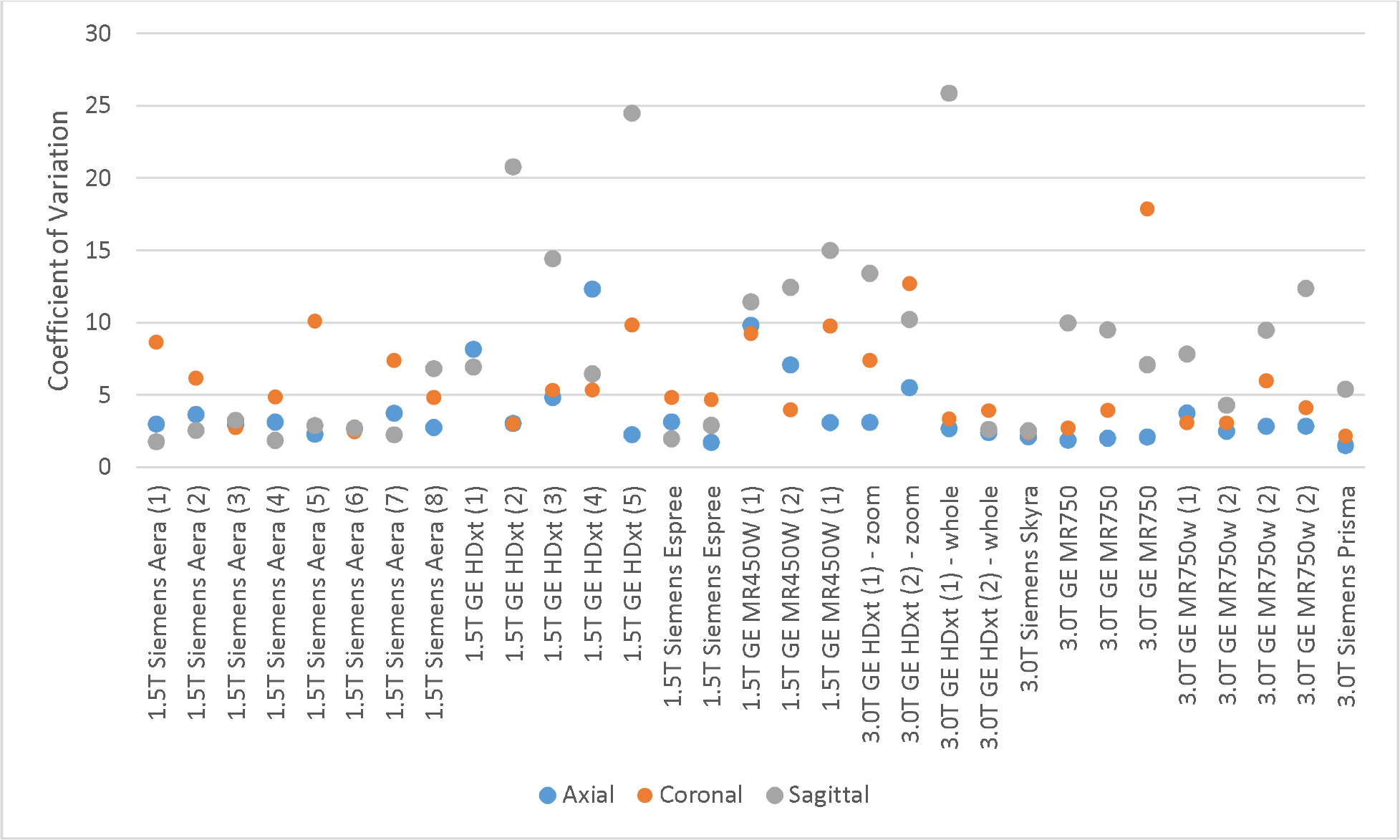
Plot of the average coefficient of variation for each MR system among the different slice orientations.

**Figure 6:**
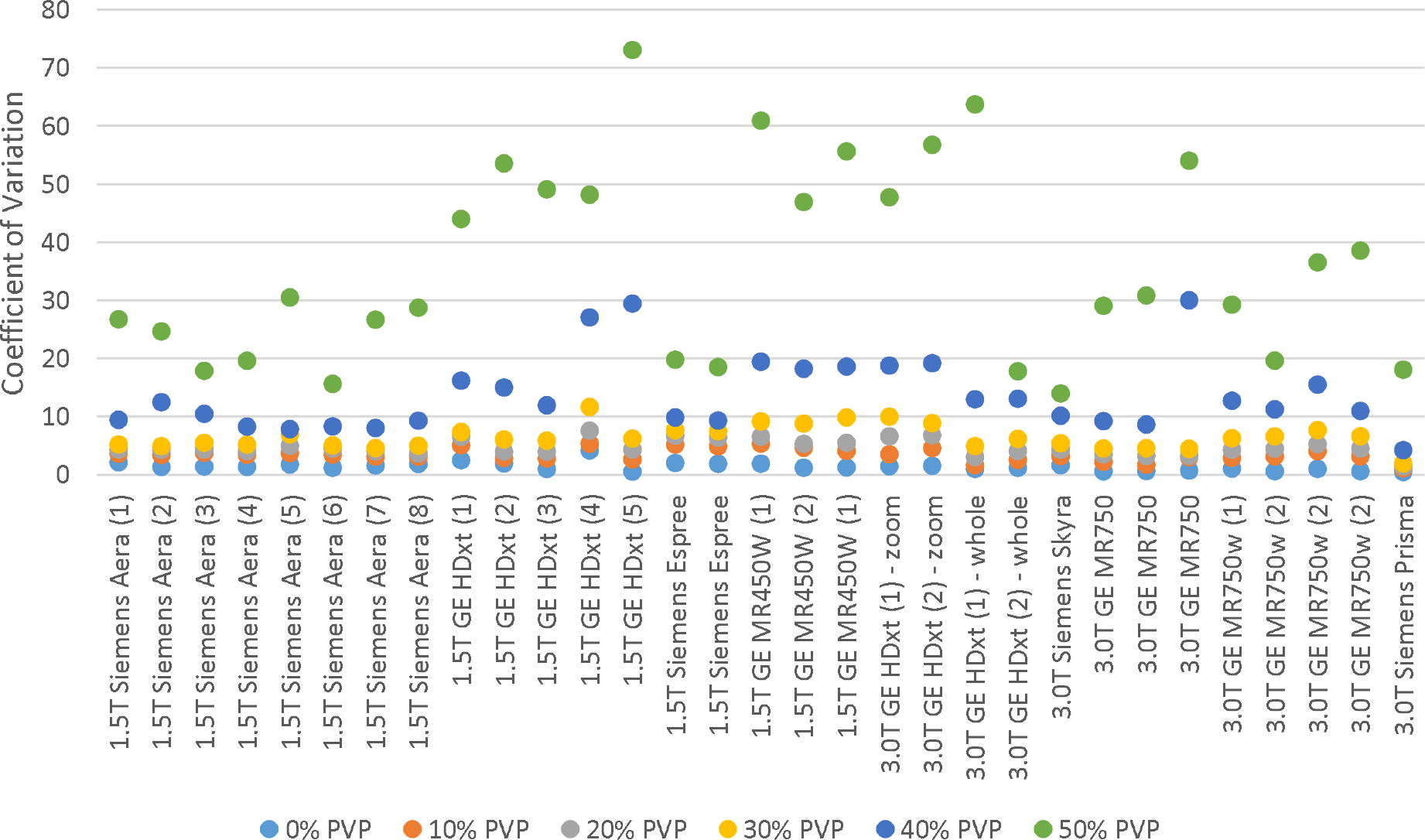
Plot of the average coefficient of variation for each MR system among the different vial PVP concentrations.

In Figures 7–9, the results from the Bland-Altman analysis are displayed. Datasets acquired from different magnetic strengths, manufacturers, and platforms were separated. The 95% limits of agreement were calculated and overlaid on to each plot, as well as the mean to represent any overall bias.

**Figure 7.**
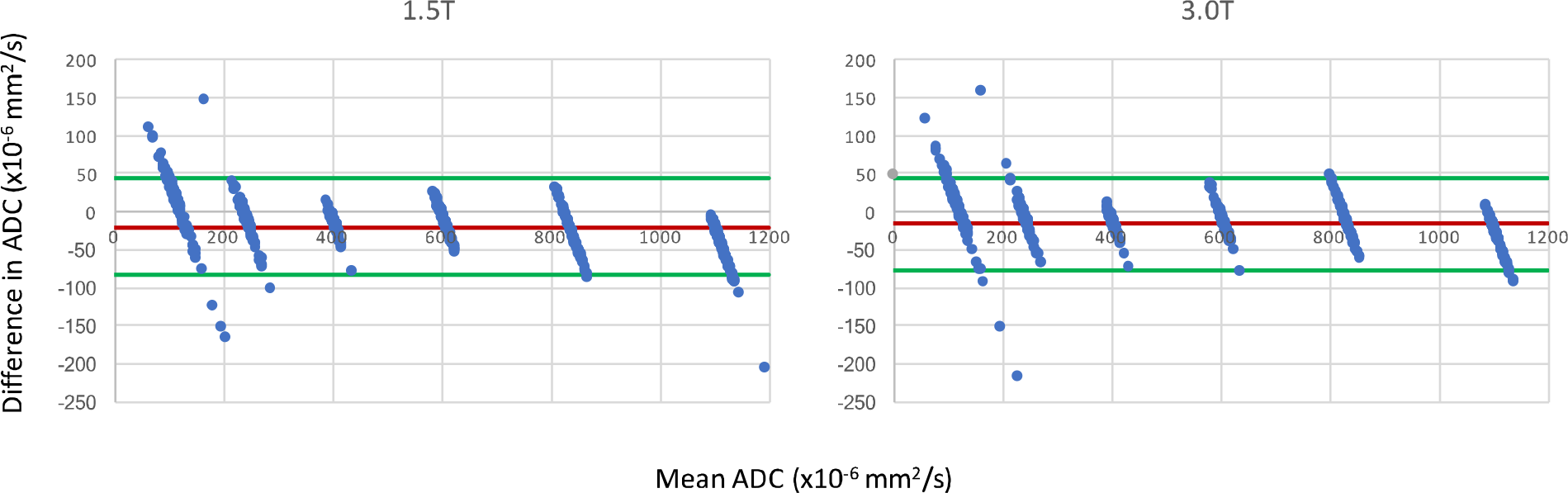
Bland-Altman plot analysis between 1.5T and 3.0T MR systems with 95% limits of agreement represented by green lines and the mean bias represented with a red line.

**Figure 8.**
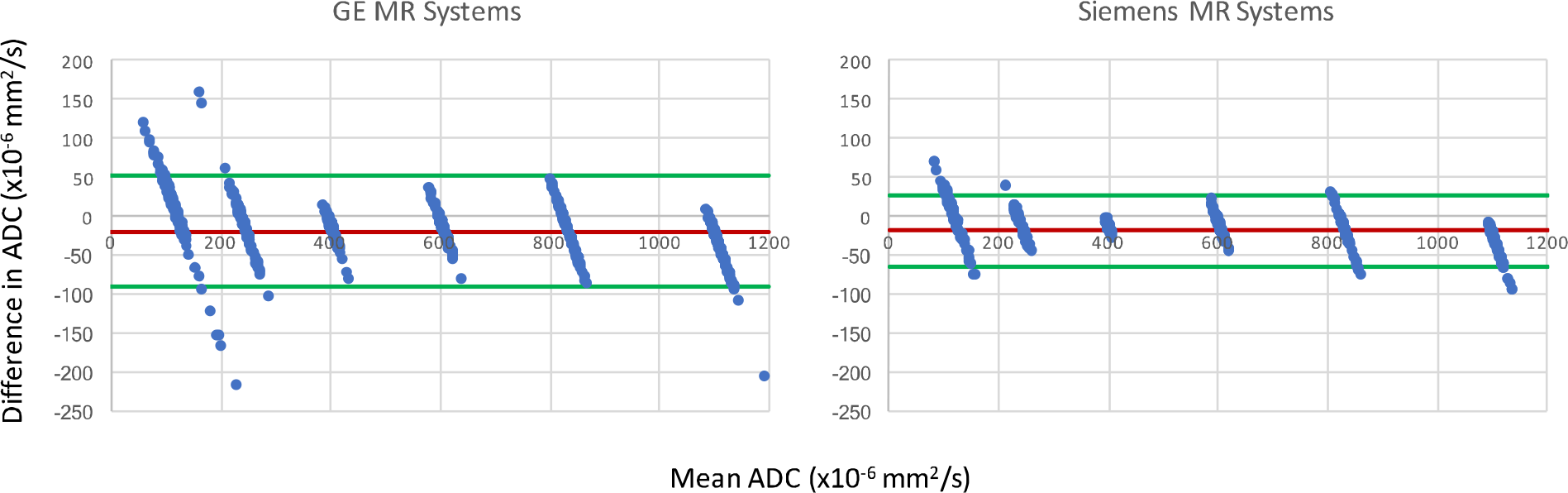
Bland-Altman plot analysis between GE and Siemens MR systems with 95% limits of agreement represented by green lines and the mean bias represented with a red line.

**Figure 9.**
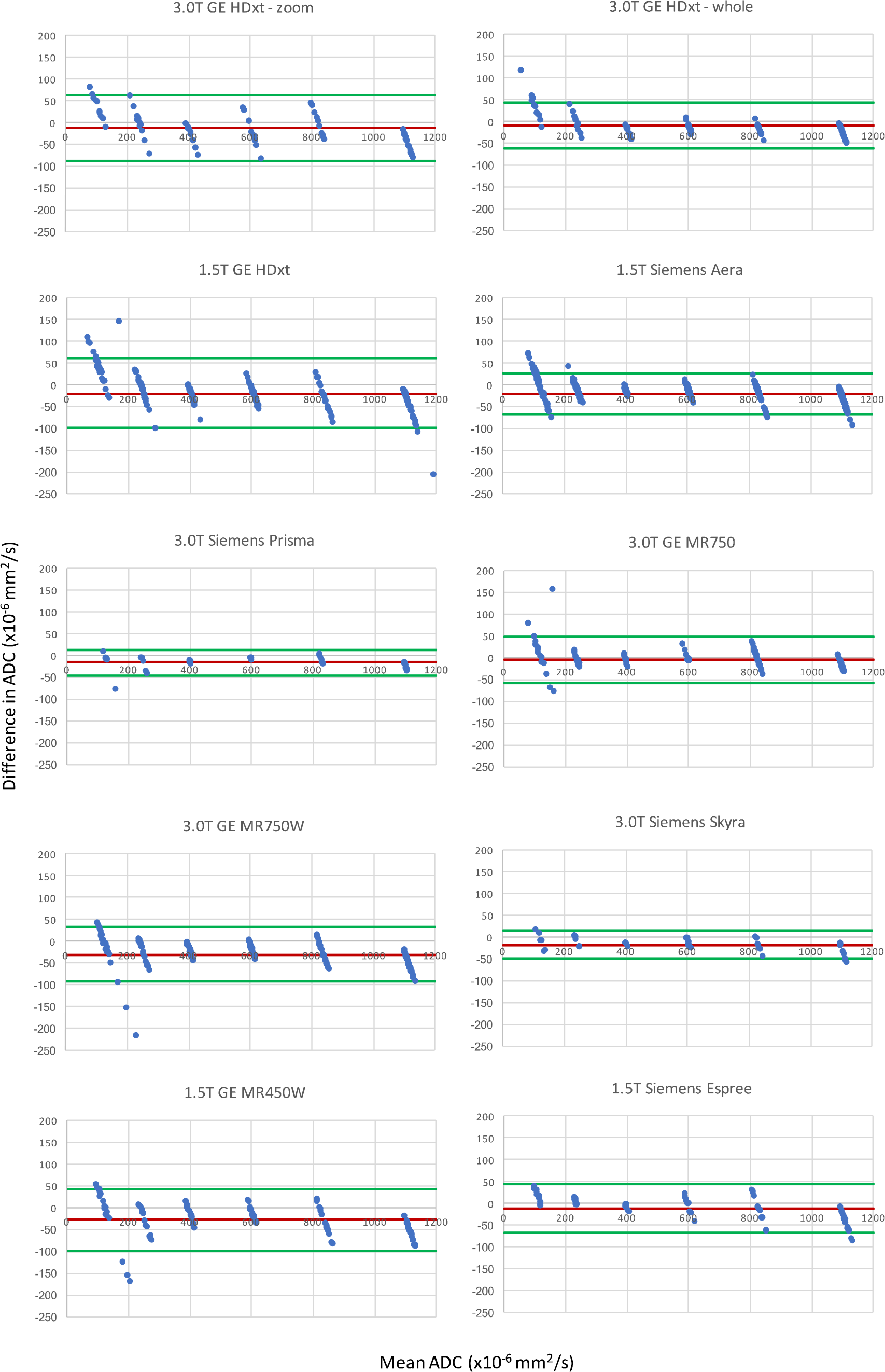
Bland-Altman plot analysis between different MR systems with 95% limits of agreement represented by green lines and the mean bias represented with a red line.

## Discussion

From the ANOVA results, the mean and standard deviation of the percent errors from each phantom were not significantly different from one another. Phantoms were used interchangeably on different scanners. From Figure 3, the large percent error seen primarily at the 50% PVP vials or low ADC values was shown to be a consequence of the SNR dependence. The large CoV in Figure 5 for the vials with the higher PVP concentration also showed them to be the least reliable.

Very little bias was seen between different magnetic strengths and manufacturers. The 95% limits of agreement were also similar between 1.5T and 3.0T acquisitions with measurements falling outside the range at the lower ADC values. Comparing between GE and Siemens MR platforms, the Siemens’ limits were narrower with less measurements outside the 95% limits. However, only 12 sets of data were acquired on Siemens MR systems compared to 19 sets on GE systems. Future work will investigate SNR dependency and compare measurements without the higher 2000 s/mm^2^ b-value.

Figure 9 showed different intervals for their respective limits of agreement. All systems seem to be close to having zero bias on average with a slightly increased error for the highly restricted, low SNR, ADC values and negative bias for high diffusion values. With the Bland-Altman analysis, it was observed that primarily lower ADC value measurements were outside the 95% limit. With the known ADC value being small, these differences produced large percent errors, compared to the same differences at the larger ADC values producing small percent errors.

## Conclusion

From this study’s acquisition and analysis of diffusion-weighted imaging data from numerous MR systems throughout the institution, the characterization of ADC variability and reliability was examined. With the increasing usage of diffusion-weighted imaging in the clinic, a quantitative quality assurance process can be implemented to provide an improved quality control over the MR systems. With more data across different MR platforms, future work would involve creating conversion factors to help facilitate quantitative measurements for multi-platform and multi-site imaging studies.

## Data Availability

Raw data are made available as a .XLS file via figshare doi with a 12-month embargo due to pending manuscript(s) under review; DICOM files are to be made publicly available after manuscript acceptance.

https://doi.org/10.6084/m9.figshare.12933938

## Acknowledgements

Dr. Fuller received/receives funding and salary support related to this project during the period of study execution from: the National Institutes of Health (NIH) National Institute of Biomedical Imaging and Bioengineering (NIBIB) Research Education Programs for Residents and Clinical Fellows Grant (R25EB025787-01); the National Institute for Dental and Craniofacial Research Establishing Outcome Measures Award (1R01DE025248/R56DE025248) and Academic Industrial Partnership Grant (R01DE028290); NCI Early Phase Clinical Trials in Imaging and Image-Guided Interventions Program (1R01CA218148); an NIH/NCI Cancer Center Support Grant (CCSG) Pilot Research Program Award from the UT MD Anderson CCSG Radiation Oncology and Cancer Imaging Program (P30CA016672); an NIH/NCI Head and Neck Specialized Programs of Research Excellence (SPORE) Developmental Research Program Award (P50 CA097007).

Dr. Fuller received/receives funding and salary support unrelated to this project during the period of study execution from: NIH Big Data to Knowledge (BD2K) Program of the National Cancer Institute (NCI) Early Stage Development of Technologies in Biomedical Computing, Informatics, and Big Data Science Award (1R01CA2148250; National Science Foundation (NSF), Division of Mathematical Sciences, Joint NIH/NSF Initiative on Quantitative Approaches to Biomedical Big Data (QuBBD) Grant (NSF 1557679); NSF Division of Civil, Mechanical, and Manufacturing Innovation (CMMI) grant (NSF 1933369); and the Sabin Family Foundation.

Direct infrastructure support is provided to Dr. Fuller by the multidisciplinary Stiefel Oropharyngeal Research Fund of the University of Texas MD Anderson Cancer Center Charles and Daneen Stiefel Center for Head and Neck Cancer and the Cancer Center Support Grant (P30CA016672) and the MD Anderson Program in Image-guided Cancer Therapy.

Dr. Fuller has received direct industry grant support, honoraria, and travel funding from Elekta AB unrelated to this project.

Part of the research was conducted at the Center for Academic Biomedical Imaging at the University of Texas MD Anderson Cancer Center with equipment support from GE Healthcare.

This research is supported in part by the National Institutes of Health through MD Anderson’s Cancer Center Support Grant CA016672. Appreciation is expressed to Dr. Thomas Chenevert for providing the analysis package used in this study.

